# Tracking the Progression & Influence of Beta-Amyloid Plaques Using Percolation Centrality and Collective Influence Algorithm: A Study using PET Images

**DOI:** 10.1101/2020.10.12.20211607

**Authors:** Gautam Kumar Baboo, Raghav Prasad, Pranav Mahajan, Veeky Baths

**Author notes:** F. author. T. author.

## Abstract

(1) Background: Network analysis allows investigators to explore the many facets of brain networks, particularly the proliferation of disease. One of the hypotheses behind the disruption in brain networks in Alzheimer’s disease is the abnormal accumulation of beta-amyloid plaques and tau protein tangles. In this study, the potential use of percolation centrality to study beta-amyloid movement was studied as a feature of given PET image-based networks; (2) Methods: The PET image-based network construction is possible using a public access database - Alzheimer’s Disease Neuroimaging Initiative, which provided 551 scans. For each image, the Julich atlas provides 121 regions of interest, which are the network nodes. Besides, using the collective influence algorithm, the influential nodes for each scan are calculated; (3) Analysis of variance (p<0.05) yields the region of interest Gray Matter Broca’s Area for PiB tracer type for five nodal metrics. In comparison, AV45: the Gray Matter Hippocampus region is significant for three of the nodal metrics. Pairwise variance analysis between the clinical groups yields five and twelve statistically significant ROIs for AV45 and PiB, capable of distinguishing between pairs of clinical conditions. Multivariate linear regression between the percolation centrality values for nodes and psychometric assessment scores reveals Mini-Mental State Examination is reliable(4) Conclusion: percolation centrality effectively (41% of ROIs) indicates that the regions of interest that are part of the memory, visual-spatial skills, and language are crucial to the percolation of beta-amyloids within the brain network to the other widely used nodal metrics. Ranking the regions of interest based on the collective influence algorithm indicates the anatomical areas strongly influencing the beta-amyloid network.

## 1. Introduction

Alzheimer’s disease predominantly stands out when it comes to neurodegenerative diseases affecting the middle-age (early-onset Alzheimer’s disease (A.D.)) and the old-age (Late-onset AD) human population. Current projections are estimated to cost about 2 trillion U.S. Dollars by 2030[1] affecting 75 million individuals by the same year. The indirect costs are estimated to be about 244 billion U.S. Dollars[2]. With no sight of a cure for A.D. and with increasing cases, early diagnosis and active management are the keys to tackling this disease for now. The ability to predict the disease’s progression with high accuracy helps design a suitable treatment regime at an early stage, thereby bringing the disease’s management to an affordable cost range.

The current methods of diagnosis of the disease include both non-invasive and invasive techniques of investigations ranging from Positron Emission Tomography (PET) scans[3] or Cerebrospinal Fluid (CSF) analysis[4]. Positron Emission Tomography or PET imaging involves the use of radiopharmaceuticals such as 2-[18F], florbetapir-fluorine-18 (AV45), or 11C-Pittsburgh compound B (PiB). AV45 and PiB[5] are comparatively newer and different in terms of the image construction mechanism. Both AV45 and PiB bind to beta-amyloid but vary in their half-life. AV45 has a half-life of 109.75 minutes and PiB, 20 minutes[6]. A comparison between PiB and AV45 varies because AV45 shows uptake within the white matter regions[7].

A combination of techniques or criteria is currently employed to detect and determine the extent of dementia due to AD. Methods which include family history, psychiatric history for cognitive and behavioral changes, which is then followed by psychometric assessments such as Mini-Mental State Examination (MMSE)[8], Frontal Assessment Battery[9] and the Neuropsychiatric Inventory Questionnaire (NPIQ)[10].

The MMSE questionnaire provides assessment in five areas of cognitive function; Orientation, Attention, Memory, Language, and Visual-Spatial skills. Similarly, the NPI questionnaire provides assessment in twelve neuropsychiatric symptoms. These two questionnaires provide the classification of the patients into three clinical conditions; cognitively normal, mild cognitive impairment, and dementia due to A.D.

The application of network analysis/graph theory to anatomical neural networks has proved useful in understanding the brain connectivity[11,12](deviations under various psychological and neurological disease states. Network analysis on neuroimaging data such as EEG, MEG, fMRI, and PET scans proves to be useful to show the variation between a cognitively normal population versus other diagnostic states using various graph-theoretic metrics[13,14].

Graph metrics such as characteristic path length, clustering coefficient, modularity, and hubs have been studied and have provided insights into the brain networks of AD patients and control groups. Some studies have tried to map the progression of MCI to dementia due to AD[15,16]; thus, network analysis and the various graph metrics have shown potential as a tool to investigate the brain networks. Network analysis on AD is a practical application wherein it describes the Alzheimer’s brain network’s behaviour. Connectivity analysis using fMRI and EEG data reports provides mixed responses; when comparing AD patients and the control group[17], there is an increase or decrease in the network’s connectivity. A reduction in connectivity could explain the cortical atrophy/disruption of the network. An increase could explain the compensatory mechanism[18].

Network Analysis on PET images related to A.D. mainly revolve around learning models or are limited to tracers that focus on the metabolic networks and the associated deviations of these networks[19,20]. Other methods include applying algorithms to the raw PET images to recognize patterns to resolve differences between healthy controls and patients with neurodegeneration[21]. The use of PET imaging and lumbar puncture to determine the levels of beta amyloids in either of them beyond the normal levels is the current standard of practice for the determination of dementia due to A.D.[22,23].

To understand beta-amyloid propagation, we propose applying graph theoretic methods on PET images to understand beta-amyloid advancement. The main benefit of adding this method is that

- This does not introduce any new steps for data collection from the patient and, at the same time.
- Adds value to the existing data by computing the percolation centrality of a given node at a given time

Network topology offers insights into the evolution of the network in a clinical setting. Studying such an evolution provides a possibility to understand the weak links within Julich atlas[24–26] based region of interest(ROI) networks. Such networks’ structural connectivity information might yield the source and sink of neurodegeneration with the brain architecture.

Percolation centrality is defined as the proportion of ‘percolated paths’ that pass through that node; this measure quantifies the relative impact of nodes based on their topological connectivity, as well as their percolated states. In other words, it is one such graph metric that looks at the extent to which a given node within a network has percolated information or can percolate information(Figure 1). The volume of information transmitted via a given node is provided by values ranging from 0.0 to 1.0[27,28]. Prior exploration of percolation centrality on disease networks[28–31] and percolation centrality in disease networks of the brain[32] have shown this as a promising metric for brain network investigation.

**Figure 1.**
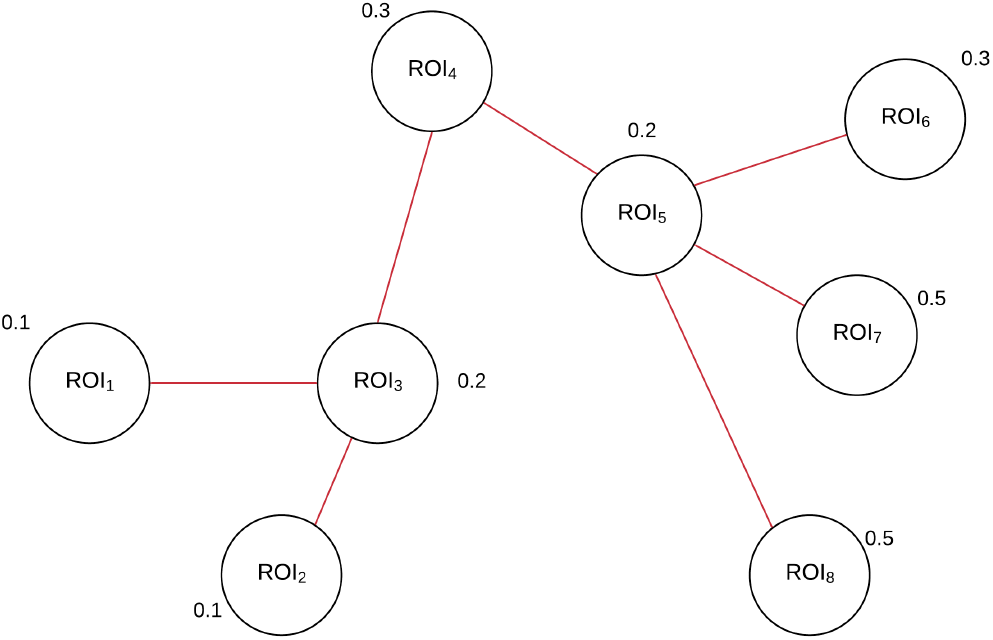
Generic example of a Percolation Network and Percolation Centrality

The knowledge on the application of percolation centrality on human PET-image-based networks is scarce at present. This work aims at adding knowledge to the gap. On the other hand, collective influence provides a minimum set of nodes or regions of the interest that can transfer information or spread disease with ease with optimal spread[33] based on the optimal percolation theory. By examining the network for the minimum set of nodes, this set will provide the regions of interest within the brain that optimally move beta-amyloid, disrupting the normal functioning of the existing neural networks.

Thus, the ability to detect the disease and predict the rate of progression of the disease at an early stage is imperative. To this end, the study aims to answer two main questions:

1. Can percolation centrality measure be used to determine the percolation of beta-amyloids within the brain?
2. Can the collective influence algorithm provide a minimum set of nodes that are vital to the AD network?

## 2. Materials and Methods

### 2.0.1. Patient Distribution

Based on the tracer agents used for acquiring the PET images, each diagnostic state subset of the data set is divided into the two available tracers; AV45[34] and PiB[6]. The patients are categorized as Cognitively normal, with Mild Cognitive Impairment, or having Alzheimer’s Disease (AD) based on the ADNI study’s psychometric assessments. Next, the PET image is matched with the patient’s diagnostic state at the time of the imaging procedure. This provided a set of observations for each type of tracer for each patient condition clinical group((Table 1). Finally, the resulting set of patients is matched with the demographic information providing 531 patients.

**Table 1:**
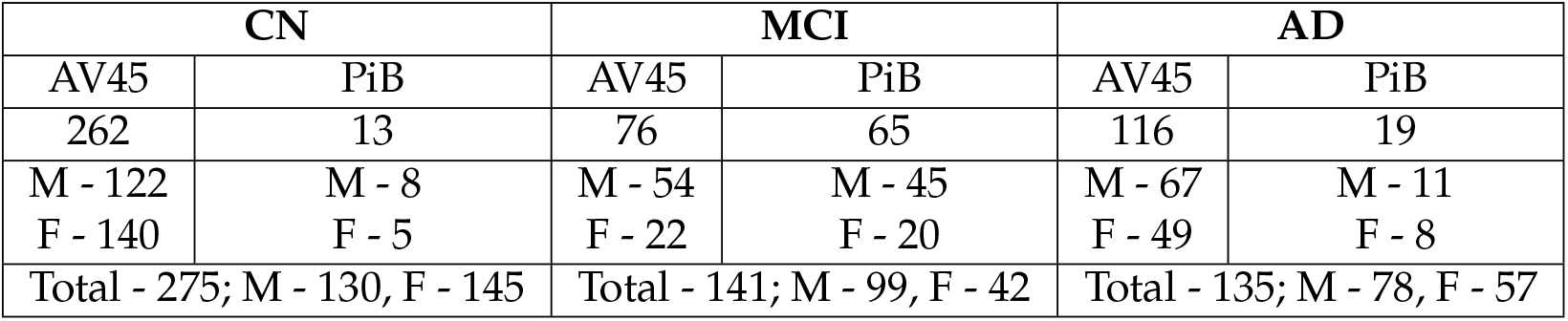
Distribution of patients

#### 2.1. Network Construction and Processing

##### 2.1.1. PET Image preprocessing

Image preprocessing is carried out in two steps (Figure 2):

**Figure 2.**
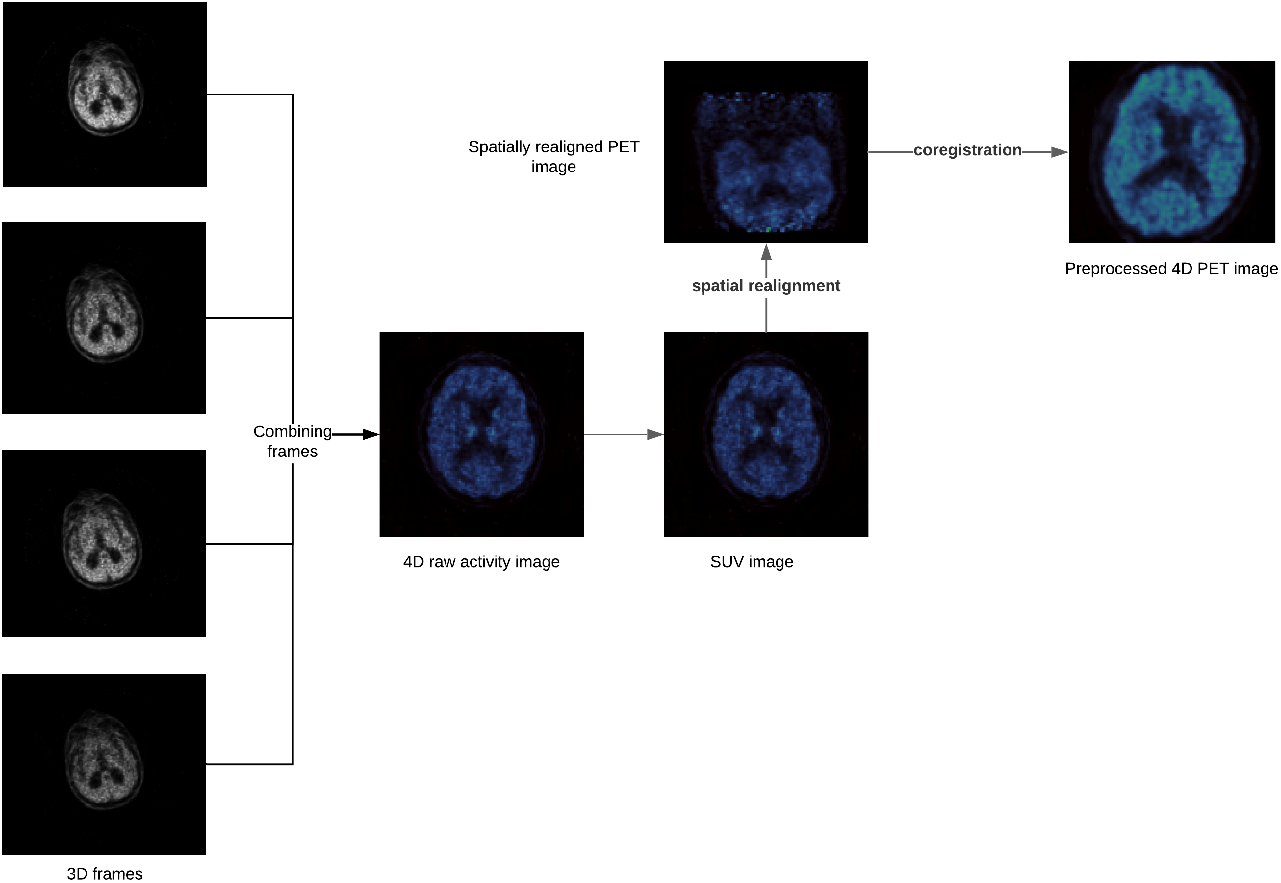
PET image preprocessing flowchart.

1. Combining individual frames of the PET image to form a 4D raw activity image. This is done using the fslmerge utility included in FSL[35].

2. The 4D raw activity image is converted to a 4D SUV image using the following formula:

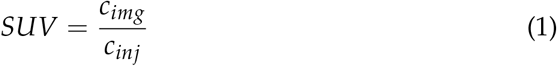

where *c*_*img*_ (Mbq ml^−1^) is given by the raw activity image and 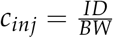 is the injection dose[36], and *BW* (g) is the body weight of the patient, considering the equivalency 1*g* = 1*ml*

3. Spatially realigning the PET frames to correct for motion. This was done using MCFLIRT.[37] The motion correction occurs with 6 DOF. The PET frames are realigned using the mean image as a template. The mean image is obtained by applying the motion correction parameters to the time series and averaging the volumes.

4. Coregistering the 4D SUV image from subject space to MNI[38] space. This is done using FreeSurfer[39]. The image used for coregistration is the MNI152_T1_2mm_brain.

To parallelize this operation, GNU Parallel[40] is used.

#### 2.2. PET Image-based Network Construction

The network is constructed using the regions of interest (ROIs) from the Julich Atlas. This atlas provides 121 ROIs, which translates to 121 nodes or vertices in the network((Figure 3). Building networks from the preprocessed images requires the generation of adjacency matrices. The adjacency matrix is computed by calculating the method described below.

**Figure 3.**
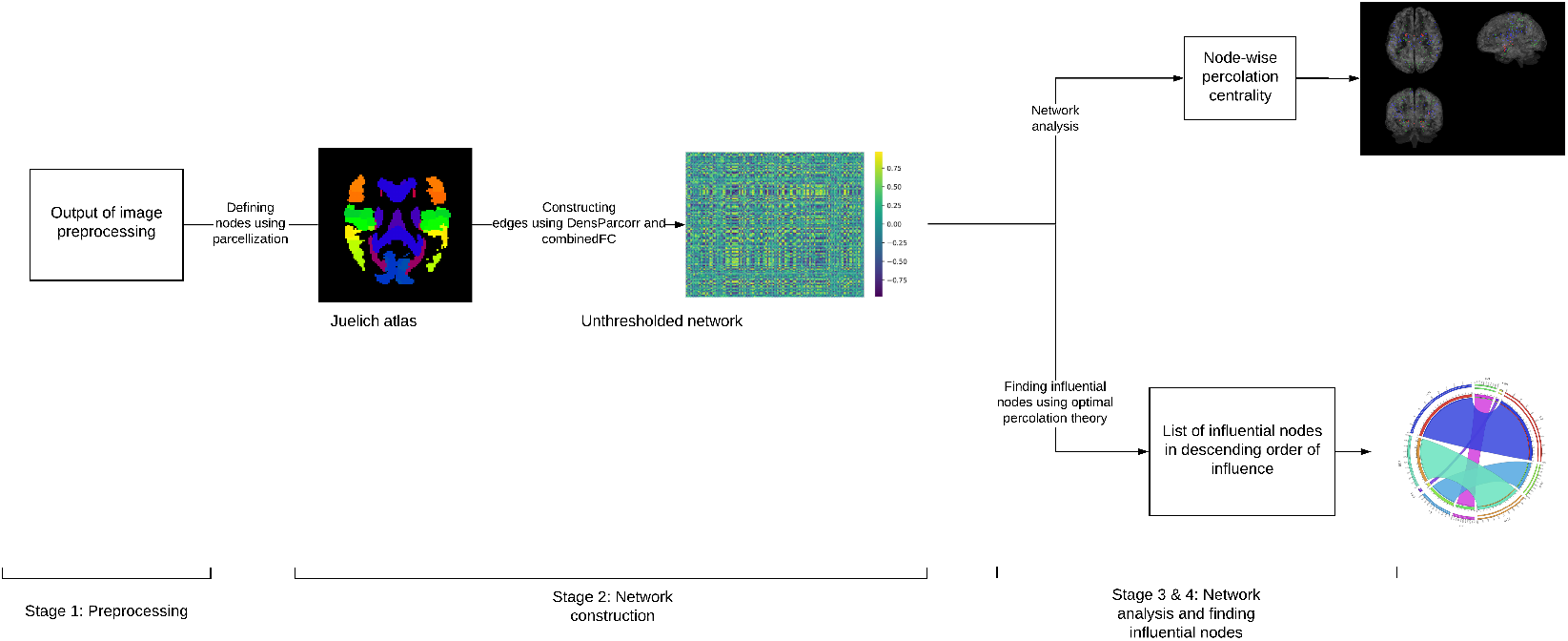
Analysis Pipeline

The Bivariate Pearson correlation performs poorly in cases of “confounding” or “chain” interactions. In such cases, partial correlation measures the direct connectivity between two nodes by estimating their correlation after regressing out effects from all the other nodes in the network, hence avoiding spurious effects in network modeling. Whereas in cases of “colliding” interactions, a partial correlation may induce a spurious correlation. Thus, Sanchez-Romero and Cole have introduced a combined multiple functional connectivity method[41].

The network is constructed by computing the pairwise partial correlation values of voxel intensities in the PET images to produce an initial adjacency matrix (*matrix*_*part*_). A second matrix (*matrix*_*bivar*_) is constructed by computing the bivariate correlation values of voxel intensities in the PET images. Now, *matrix*_*part*_ is modified using *matrix*_*bivar*_ as follows:

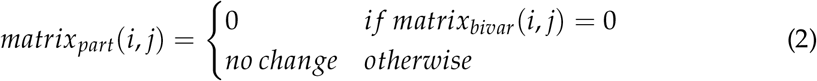

where *matrix*_*part*_(*i, j*) and *matrix*_*bivar*_(*i, j*) is the element at (*i, j*) in the respective matrices. *matrix*_*part*_ is now the combinedFC adjacency matrix that defines the network((Figure 4).

**Figure 4.**
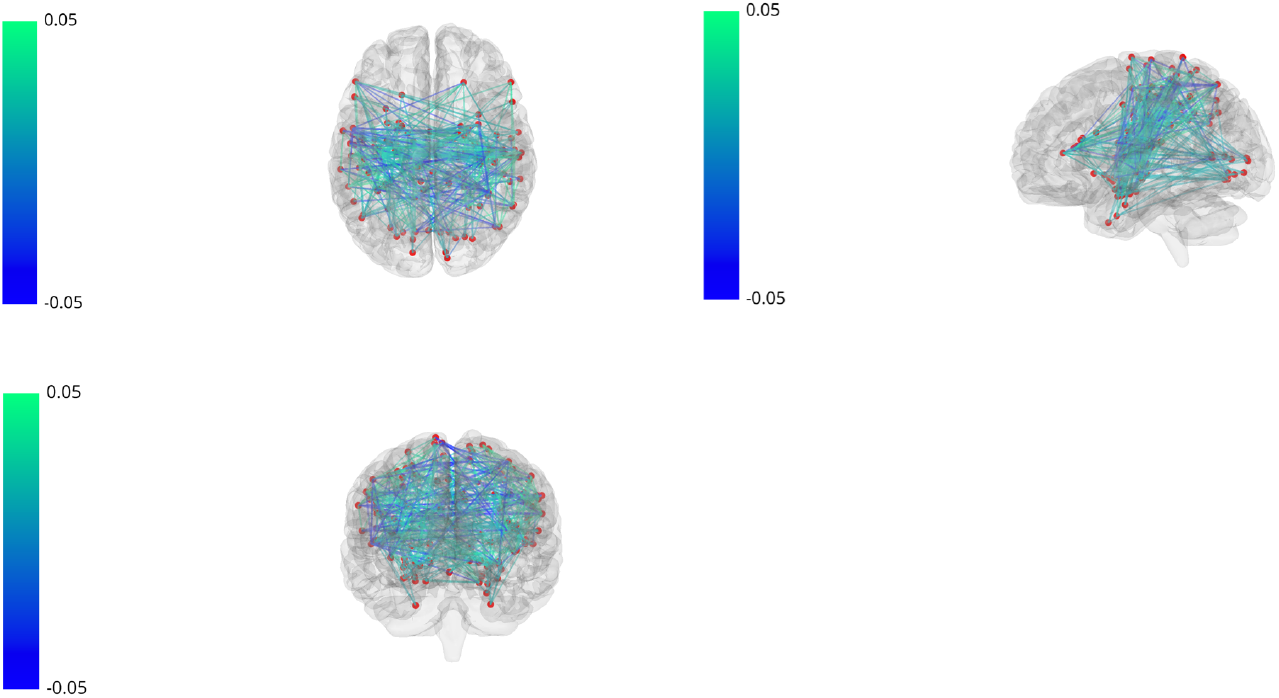
A connected network of all the nodes using the Julich Atlas. Green circles indicate the ROIs, the connecting lines indicate the edges with their weights as denoted by the accompanying color bar

The partial correlation is calculated using the correlation between two residuals; the values are computed using *N* − 2 ROIs as co-factors for every pair of ROIs[42]. The partial correlation values serve as the edge weights and constitute the values in the adjacency matrices.

Partial correlations are computed as correlation of residuals. The first order partial correlation (*ρ*_*ij*.*k*_) of *x*_*i*_ and *x*_*j*_, controlling for *x*_*k*_ is given by [43].

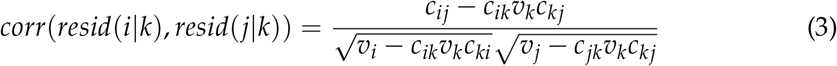

where *c*_*ij*_ = *cov*(*x*_*i*_, *x*_*j*_) and *v*_*k*_ = *var*(*x*_*k*_) Further,

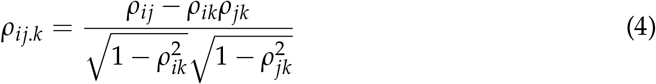

Since we are controlling for (*N* − 2) ROIs for each pair of ROIs *ROI*_*i*_ and *ROI*_*j*_, we calculate the (*N* − 2)^*th*^ order partial correlation. This is calculated recursively as

*For each ROI*_*k*_ ∈ **ROIs**

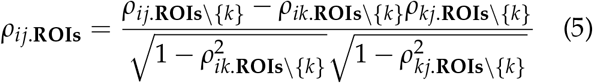

The base case of this recursive algorithm is given by equation 3. The estimation of partial correlations is a computationally intensive task, mainly due to the pre-calculation of residuals before computing cross-correlation. And because the number of covariates is large; this calculation is done in a time-optimized manner using the R package ppcor[43]. Next, the adjacency matrices with set threshold; using a) data-driven threshold scheme based on Orthogonal Minimal Spanning Trees (OMSTs)[21,44,45]. Network threshold serves to remove inconsequential (or low-impact) edges and reduce the network complexity. And b) shortest path thresholding scheme [46]. This is done to compare between the two common schemes used for threshold.

The Networkx[47] Python library is used for network construction from the threshold adjacency matrices and subsequent percolation centrality computation and other graph metrics.

#### 2.3. Percolation Centrality Computation

Percolation centrality is a nodal metric and is calculated for each node. The percolation centrality for each node v at time t is calculated as shown below:

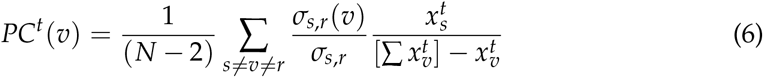

Where *σ*_*s,r*_ is the number of shortest paths between nodes *s* and *r* pass-through node *v*,

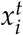 is the percolation state of node *i* at time *t*,

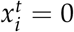 = 0 indicates a non-percolated node and,

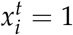 = 1 indicates a fully percolated node.

The percolation centrality value is calculated for each network using the inbuilt function of Networkx.(see supplementary data)

#### 2.4. Collective Influence Algorithm

We define *G*(*q*) the fraction of occupied sites (or nodes) belonging to the giant(largest) connected component. Percolation theory[48] tells us that if we choose these *q* fraction of nodes randomly, the network undergoes a structural collapse at a certain critical fraction where the probability of existence of the giant connected component vanishes, *G* = 0. The optimal percolation problem is finding the minimum fraction *q*_*c*_ of nodes to be removed such that *G*(*q*_*c*_) = 0 i.e. the minimum fraction of “influencers” to fragment the network. For any fixed fraction *q < q*_*c*_, we search for the configuration of removed nodes that provides the minimal non-zero giant connected component *G*. For further reading on how the problems of optimal immunization and spreading (optimal influencer problem) to the problem of minimizing the giant component of a network, i.e., optimal percolation problem, readers are encouraged to read [49].

The algorithm is on the basis that, given a network: the flow of information within the network is optimal with a minimum number of nodes that weigh heavily on the flow of information through the said network[33]. In the context of this investigation, the small sets of nodes/ROIs would prove to be vital in the movement of beta-amyloid plaques.

The core idea is that the overall functioning of a network in terms of the spread of information (or in our case, movement of beta-amyloid plaques) hinges on a specific set of nodes called influencers. This idea of finding the most influential nodes has been previously used in other contexts, for example, activating influential nodes in social networks to spread information[50] or de-activating or immunizing influential nodes to prevent large scale pandemics[30,51]. In recent applications to neuroscience, this method has been used to find nodes essential for global integration of a memory network in rodents[32]. Our work is the first to apply it to study the progression of AD, to the best of our knowledge. In the context of this investigation, these small sets of influential nodes/ROIs would prove to be vital in the movement of beta-amyloid plaques.

With the implementation of Collective Influence (CI) algorithm, it facilitates to pinpoint the most influential nodes, more efficiently than previously known heuristic techniques. CI is an optimization algorithm that aims to find the minimal set of nodes that could fragment the network in optimal percolation, or in a sense, their removal would dismantle the network in many disconnected and non-extensive components. In percolation theory, if we remove nodes randomly, the network would undergo a structural collapse at a critical fraction where the probability that the giant connected component exists is *G* = 0. The optimal percolation is an optimization problem that attempts to find the minimal fraction of influencers *q* to achieve the result *G*(*q*) = 0.

#### 2.5. Other Graph metrics

Besides the percolation centrality measure, four other nodal metrics of a graph are calculated here. Below are the four metrics that are computed for the PET image based graphs.

##### 2.5.1. Betweenness Centrality

The basic definition of betweenness centrality is defined as,

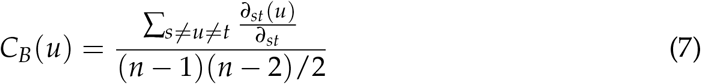

This centrality information provides the uniqueness of it within the network. In this study it provides the regions of interest that play a vital role in the information flow in the network; the information being beta-amyloids or tau proteins accumulation in those regions.

##### 2.5.2. Closeness Centrality

The closeness centrality of a node denotes how close a node is in the given network. It is inversely proportional to the farness of the node. Freeman defined the closeness centrality as,

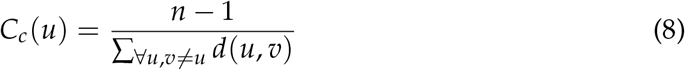

The distance between two nodes/ROIs u and v in a network, denoted d(u,v), is defined as the number of hops made along the shortest path between u and v. In this case, lesser the hops; closer are the two ROI’s and the ease with which the misformed proteins can travel.

##### 2.5.3. Current Flow Betweenness Centrality

Given a source ROI(*s*) and a target ROI(*t*), the absolute current flow through edge (*i, j*) is the quantity 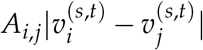. By Kirchhoff’s law the current that enters a node is equal to the current that leaves the node. Hence, the current flow 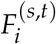 through a node *I* different from the source *s* and a target *t* is half of the absolute flow on the edges incident in *i*:

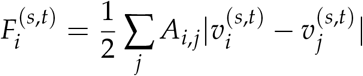

Moreover, the current flows 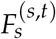 and 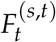 through both *s* and *t* are set to 1, if end-points of a path are considered part of the path, or to 0 otherwise. Since the potential 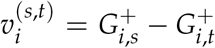, with *G*^+^ the generalized inverse of the graph Laplacian, the above equation can be expressed in terms of elements of *G*^+^ as follows:

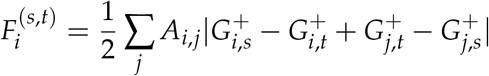

Finally, the current-flow betweenness centrality *b*_*i*_ of node *i* is the flow through *i* averaged over all source-target pairs (*s, t*):

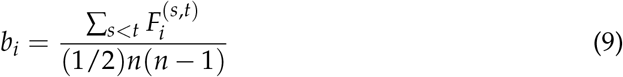

##### 2.5.4. Eigenvector Centrality

Eigenvector centrality is a measure of the influence a node has on a network. If a node is pointed to by many nodes (which also have high Eigenvector centrality) then that node will have high Eigenvector centrality. In this case, the ROI’s are the nodes and PET images provide the intensity value for each ROI, which helps in computing the centrality values each scan. Another interpretation to the centrality measure is that it provides the list of prominent regions in the brain network hierarchy and helps in detection of localized differences between patient populations[52].

Previous work on AD patients and comparison with normal patients provides the usefulness of Eigenvector centrality[53].

## 3. Statistical Analysis

For this study the null hypothesis is that percolation centrality value does not indicate the propagation of beta-amyloids within the brain network.

To determine the impact the percolation value has over each PET scan, a comparison with the regions of interest from the brain atlas is done using the Multiple linear regression analysis.

This study is exploratory in nature, and that the multiplicity problem is significant. And implementation of multiple test procedures does not solve the problem of making valid statistical inference for hypotheses that were generated by the data. But it does assist in describing the possible mechanism.

### 3.1. Pairwise Analysis of Variance

To obtain pairwise group differences, we carry out a post prior (post hoc) analysis using scikit-posthocs package; the Student T-test pairwise gives us the respective p values. The ANOVA test is performed for each node in the network with the null hypothesis that the mean percolation centrality of that node is the same across the three stages. To test the null hypothesis, Analysis of variance with significance level (*α*) of 0.05 is used.

### 3.2. Error Correction

To control for multiple comparisons of 121 nodes, the Scheffe Test and control for Experiment-wise Error Rate (EER) is carried out. It is a single-step procedure that calculates the simultaneous confidence intervals for all pairwise differences between means.

### 3.3. Multivariate Linear Regression

A correlation between the percolation centrality values for all 121 nodes and psychometric test scores - MMSE and NPIQ - is computed to identify the regions of interest that can be used as reliable predictors. Instead of performing multiple correlations across all three diagnoses, a multivariate regression analysis using regularisation techniques, wherein the features are the nodal percolation centrality values, and the target variable is the MMSE or NPIQ score. The goal is not to build a predictive model but to use it to quantify each node’s influence in distinguishing between the clinical conditions for interpretation purposes. Had the purpose been building a machine learning model, it would imply the need to develop elaborate features sets (more than just percolation centrality) and utilize complex machine learning architectures (which provide less room for interpretability)

### 3.4. Regularization and Cross-Validation

We use regularization in our multivariate linear regression (MLR) to make sure our regression model generalizes better to unseen data. Regularization is necessary to control for overfitting. Here, both Lasso regression (L1 penalty) as well as Ridge regression (L2 penalty) are tested, and both provide similar root mean squared errors (RMSE) and similar desired results. We choose Lasso with *α* = 0.1, for reporting our results (Figure 6). To quantify the robustness and reliability of our model, before and after regularization, we perform a leave one out cross-validation (LOOCV). We choose this cross-validation strategy because it is unbiased and better suited to our smaller sample sizes (especially in PiB tracer subset). We observe an improvement in validation RMSE with an increase in regularization (parameter *α*), but we also observe that excessive penalization of weights at very high values of *α* can result in the regression model converging to the mean of the output MMSE/NPIQ scores. To take this into account, we also plot the standard deviation in predicted MMSE/NPIQ outputs and choose *α* = 1 for sufficient but not excessive regularization (Further details in supplementary figures).

**Figure 5.**
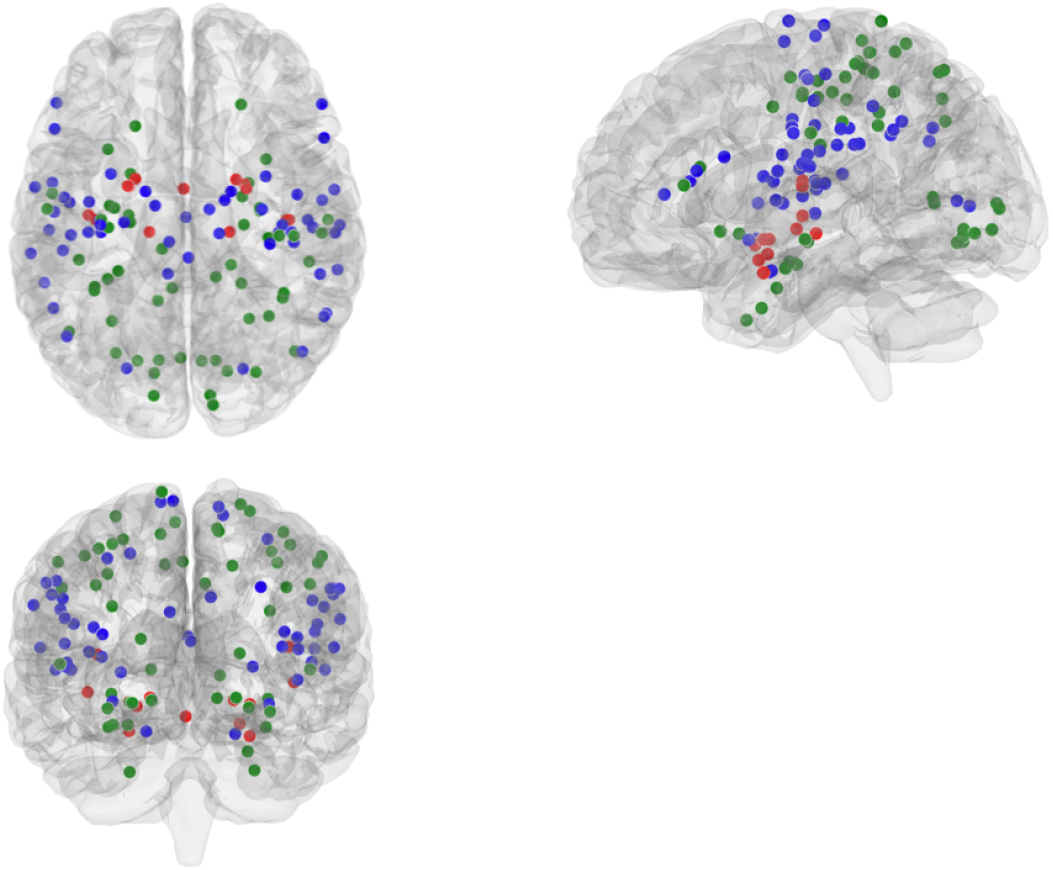
Illustrates the ROIs that corresponds to MMSE and NPIQ. The green circles represent ROIs associated with the MMSE psychometric assessment, the red circles represent ROIs associated with the NPIQ psychometric assessment, and the blue circles represent ROIs associated with both MMSE and NPIQ

**Figure 6.**
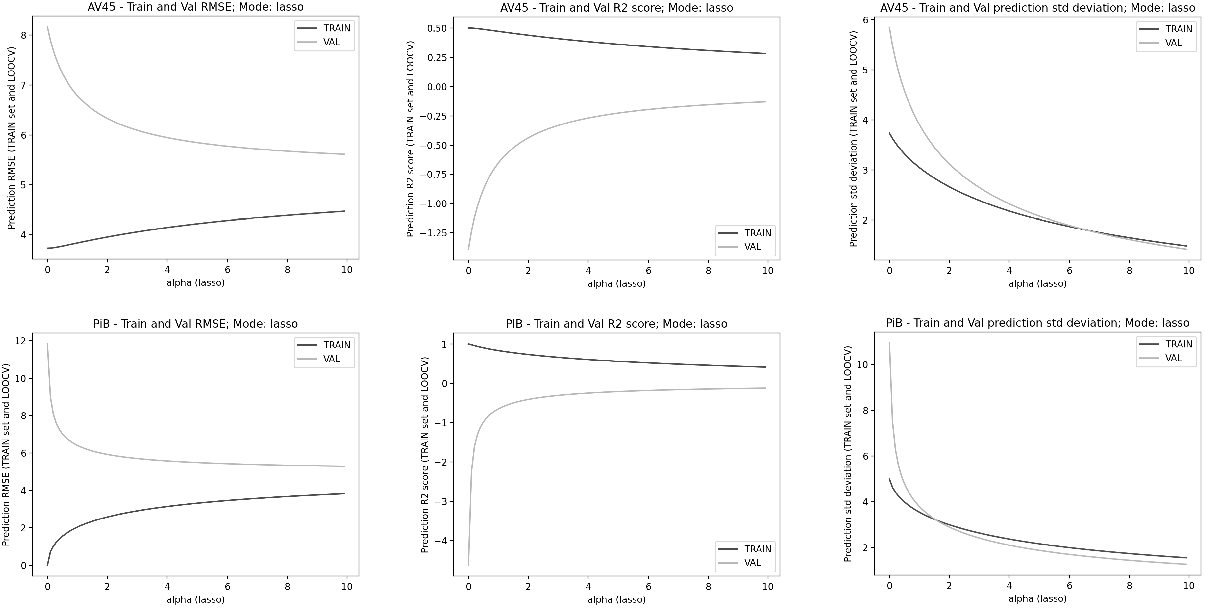
Regularization using Lasso regression with L1 penalty.

## 4. Results

### 4.1. Pairwise ANOVA

The student t-test is performed on the resulting five centrality values for each tracer type and clinical conditions. There was a significant effect of the beta-amyloid accumulation on the five centrality values at p<0.05 level for the three clinical groups[F(3, 454) = 3.002 for AV45 and F(3, 97) = 3.027 for PiB] (Table 2). A one-way between clinical groups ANOVA was conducted to compare the effect of beta-amyloid accumulation/tau protein on five centrality values in the cognitive normal, mild cognitive impairment and Alzheimer’s disease patient(Table 3 & Table 4)).

**Table 2:** Number of Scans per tracer type and corresponding critical F-values

| Tracer | AV45 | PiB |
| --- | --- | --- |
| No. of scans | 454 | 97 |
| Critical F-value | 3.002 | 3.027 |

CN: Cognitively Normal, MCI: Mild Cognitive Impairment, AD: Alzheimer's Disease, M: Male, F: Female

**Table 3:** Pairwise ANOVA for AV45 & PiB tracers for Betweenness Centrality

| Tracer Type | AV45 |  |  | PiB |  |  |
| --- | --- | --- | --- | --- | --- | --- |
| Thresholding scheme | ROI | f-value | p-value | ROI | f-value | p-value |
| OMST | GM Premotor cortex BA6 R | 5.94059 | 0.00283 | GM Broca's area BA45 R | 5.17524 | 0.00738 |
|  | GM Hippocampus subiculum L | 4.96072 | 0.00738 | GM Broca's area BA45 L | 4.75064 | 0.01083 |
|  | GM Visual cortex V5 L | 4.39253 | 0.01288 | GM Anterior intraparietal sulcus hIP3 R | 3.79312 | 0.02605 |
|  | GM Superior parietal lobule 7M L | 4.34316 | 0.01352 | GM Broca's area BA44 L | 3.41149 | 0.03713 |
|  | GM Primary motor cortex BA4a L | 3.73311 | 0.02463 | GM Mammillary body | 3.28404 | 0.04182 |
|  | GM Primary somatosensory cortex BA1 R | 3.56446 | 0.02908 |  |  |  |
|  | GM Primary auditory cortex TE1.2 R | 3.31334 | 0.03725 |  |  |  |
| SPT | GM Insula Ig2 L | 4.58607 | 0.01065 | GM Broca's area BA44 R | 6.22323 | 0.00290 |
|  | WM Acoustic radiation R | 4.20148 | 0.01554 | GM Primary auditory cortex TE1.2 R | 5.14313 | 0.00759 |
|  | GM Hippocampus subiculum L | 3.89671 | 0.02097 | WM Cingulum R | 4.61277 | 0.01227 |
|  | GM Mammillary body | 3.63729 | 0.02707 | GM Inferior parietal lobule PFcm R | 3.96185 | 0.02229 |
|  | GM Inferior parietal lobule Pga R | 3.04114 | 0.04872 |  |  |  |

**Table 4:** Pairwise ANOVA for AV45 & PiB tracers for Percolation Centrality

| Tracer Type | AV45 |  |  | PiB |  |  |
| --- | --- | --- | --- | --- | --- | --- |
| Thresholding scheme | ROI | f-value | p-value | ROI | f-value | p-value |
| OMST | GM Premotor cortex BA6 R | 5.87528 | 0.00302 | GM Broca's area BA45 R | 5.14527 | 0.00758 |
|  | GM Hippocampus subiculum L | 4.85879 | 0.00815 | GM Broca's area BA45 L | 4.82934 | 0.01008 |
|  | GM Superior parietal lobule 7M L | 4.48843 | 0.01172 | GM Anterior intra-parietal sulcus hIP3 R | 3.87541 | 0.02414 |
|  | GM Visual cortex V5 L | 4.38407 | 0.01299 | GM Broca's area BA44 L | 3.42666 | 0.03661 |
|  | GM Anterior intra-parietal sulcus hIP1 L | 4.14044 | 0.01650 | GM Mamilary body | 3.33030 | 0.04005 |
|  | GM Superior parietal lobule 7P R | 3.83170 | 0.02235 |  |  |  |
|  | GM Primary motor cortex BA4a L | 3.77499 | 0.02364 |  |  |  |
|  | GM Primary somatosensory cortex BA1 R | 3.65131 | 0.02670 |  |  |  |
|  | GM Primary auditory cortex TE1.2 R | 3.34477 | 0.03611 |  |  |  |
| SPT | GM Insula Ig2 L | 4.64660 | 0.01004 | GM Broca's area BA44 R | 6.30152 | 0.00270 |
|  | WM Acoustic radiation R | 4.38388 | 0.01299 | GM Primary auditory cortex TE1.2 R | 5.03631 | 0.00836 |
|  | GM Hippocampus subiculum L | 3.93752 | 0.02014 | WM Cingulum R | 4.57068 | 0.01275 |
|  | GM Mamilary body | 3.54618 | 0.02961 | GM Inferior parietal lobule PFcm R | 3.94533 | 0.02263 |
|  | GM Inferior parietal lobule Pga R | 3.17572 | 0.04266 |  |  |  |
|  | GM Lateral geniculate body L | 3.05960 | 0.04784 |  |  |  |

#### 4.1.1. Cross-Validation and Regularization

Increasing regularization (*α*) improves the validation RMSE, making it more robust and generalize to unseen data. But at higher values of *α*, it is observed that the standard deviation of predicted MMSE scores decreases to less than < 2, irrespective of clinical condition. Which could mean that it saturates to predicting the mean MMSE value when regression weights are extremely penalized. Thereby choosing a reasonably small yet effective *α* value (less than 2), for which the validation RMSE and the standard deviation in output predicted MMSE.

### 4.2. Multivariate Linear regression

A linear regression model between the percolation centrality values for all 121 nodes and psychometric test scores - MMSE and NPIQ - is computed to identify the regions of interests that can be used as reliable predictors(Figure 4). Instead of performing multiple correlations across all three diagnoses, a multivariate regression analysis using leave one out cross validation is carried out, wherein the features are the nodal percolation centrality values and the target variable are the psychological assessment scores(Table 6 & Table 7).

**Table 5:**
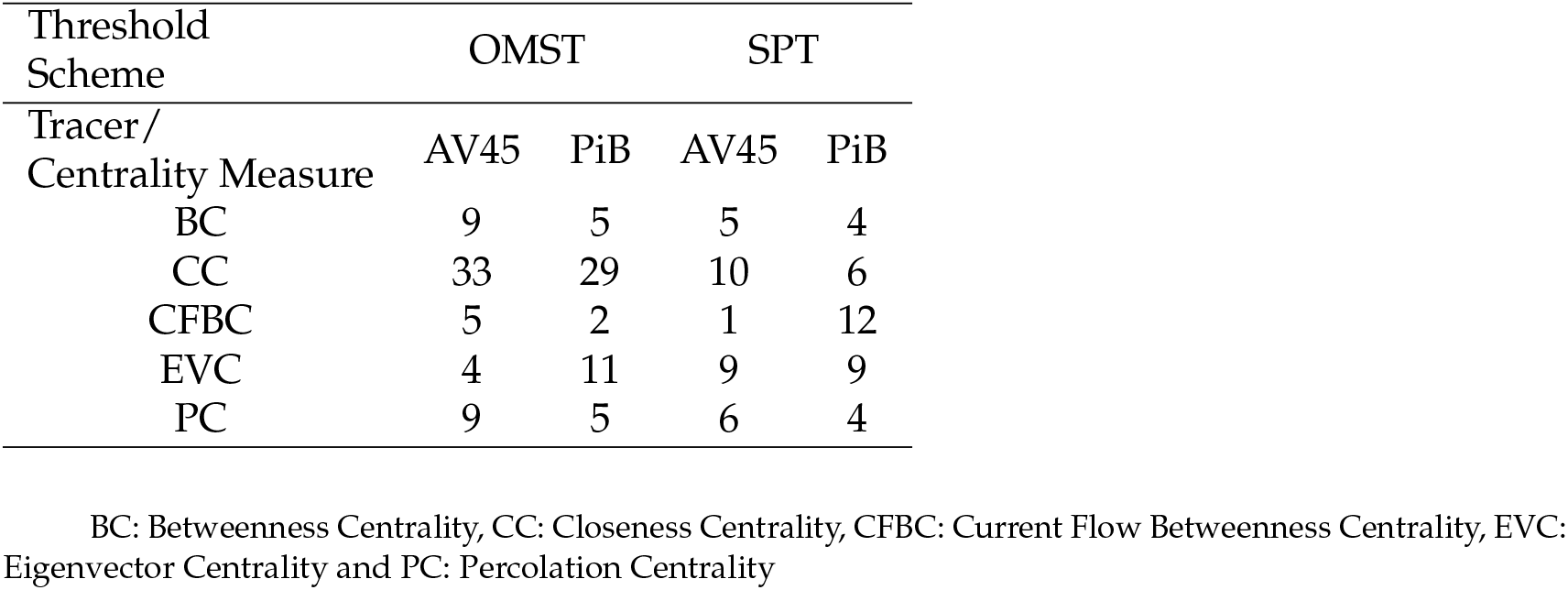
Distribution of ROI’s across graph metrics and tracer type on the basis of ANOVA test.

**Table 6:** Multivariate Linear Regression Analysis-Number of Region of Interest Across Clinical Conditions for Both Threshold Schemes for Betweenness Centrality

| Clinical Condition |  |  | CN |  | MCI |  | AD |  |
| --- | --- | --- | --- | --- | --- | --- | --- | --- |
| Graph Metric | PT | Tracer | OMST | SPT | OMST | SPT | OMST | SPT |
| Betweenness Centrality | MMSE | AV45 | 4 | 5 | 13 | 8 | 4 | 7 |
|  |  | PiB | 5 | 4 | 5 | 1 | 2 | 9 |
|  | NPI-Q | AV45 | 0 | 3 | 1 | 1 | 0 | 0 |
|  |  | PiB | 2 | 14 | 1 | 0 | 3 | 10 |

**Table 7:**
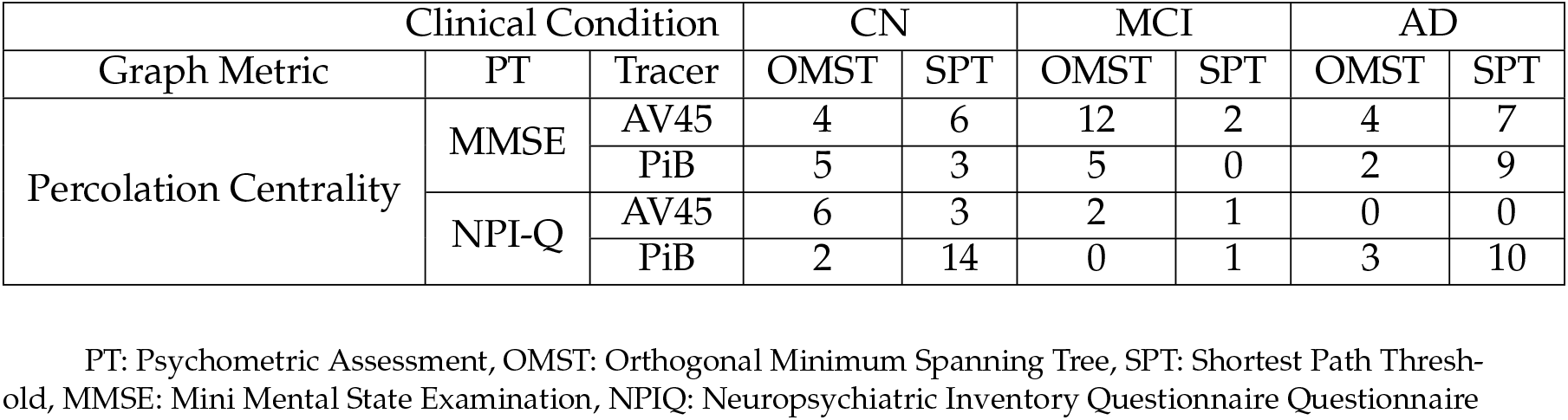
Multivariate Linear Regression Analysis-Number of Region of Interest Across Clinical Conditions for Both Threshold Schemes for Percolation Centrality

| Clinical Condition |  |  | CN |  | MCI |  | AD |  |
| --- | --- | --- | --- | --- | --- | --- | --- | --- |
| Graph Metric | PT | Tracer | OMST | SPT | OMST | SPT | OMST | SPT |
| Percolation Centrality | MMSE | AV45 | 4 | 6 | 12 | 2 | 4 | 7 |
|  |  | PiB | 5 | 3 | 5 | 0 | 2 | 9 |
|  | NPI-Q | AV45 | 6 | 3 | 2 | 1 | 0 | 0 |
|  |  | PiB | 2 | 14 | 0 | 1 | 3 | 10 |

PT: Psychometric Assessment, OMST: Orthogonal Minimum Spanning Tree, SPT: Shortest Path Threshold, MMSE: Mini Mental State Examination, NPIQ: Neuropsychiatric Inventory Questionnaire

### 4.3. Comparison of Threshold Schemes

The two schemes are compared on the number of ROI’s that can be considered on the basis of ANOVA analysis(p ≤ 0.05), The Juelich atlas has five clusters; frontal, parietal, temporal, occipital lobes and the white matter regions. As well as comparing the performance of the threshold schemes between the two tracers(Table 5). Orthogonal Minimum Spanning Tree: provides a total of 112 ROI’s across the five nodal metrics. On the Basis of the tracers, 60 ROI’s are obtained for AV45 and 52 for PiB. Further, the ranking of ROI on the basis of the threshold scheme is listed for the three clinical groups for the respective tracers

Shortest Path Threshold: here a total of 66 ROI’s are obtained. 31 and 35 for AV45 and PiB tracers respectively. On the basis of the collective influence algorithm the ROI’s are ranked for the three clinical groups and their respective tracers.

Further, comparing the number of statistically valid(MLR) ROIs across the five centrality values with the psychometric tests; MMSE and NPI-Q is performed. This helps in comparing the function of the ROI with the assessment carried out on the test(Table 5 Also see supplementary material-Tables 11 through 18).

### 4.4. Other Graph Metrics

The Closeness Centrality provides the highest number of ROI’s across overall(Table 10); 80 in total. Eigenvector Centrality provides 33 ROI’s(Table 9), whereas Percolation Centrality has 24 ROI’s followed by the Betweenness Centrality measure with a total of 23 ROI’s and Current Flow Betweenness Centrality 19 ROI’s across the two tracers(Table 8).

**Table 8:**
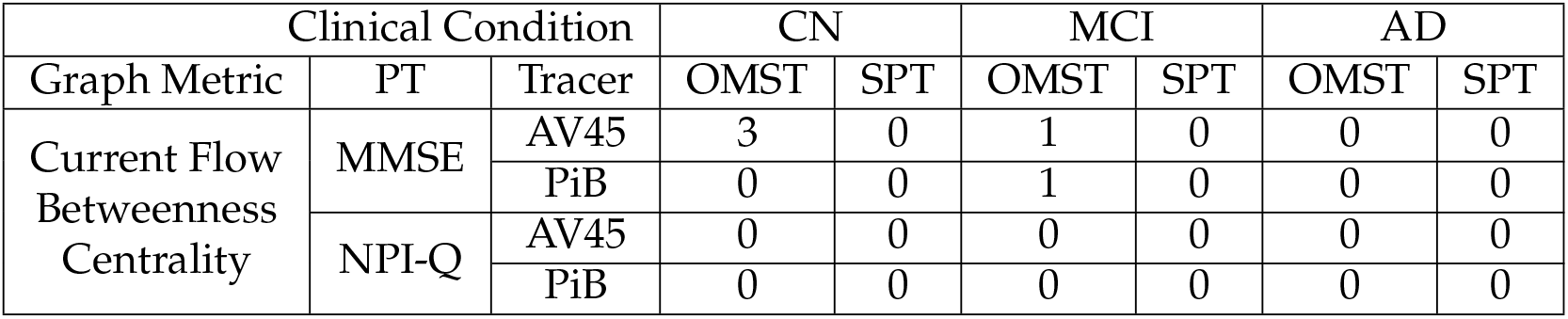
Multivariate Linear Regression Analysis-Region of Interest Across Clinical Conditions for Both Threshold Schemes for Current Flow Betweenness Centrality

| Clinical Condition |  |  | CN |  | MCI |  | AD |  |
| --- | --- | --- | --- | --- | --- | --- | --- | --- |
| Graph Metric | PT | Tracer | OMST | SPT | OMST | SPT | OMST | SPT |
| Current Flow Betweenness Centrality | MMSE | AV45 | 3 | 0 | 1 | 0 | 0 | 0 |
|  |  | PiB | 0 | 0 | 1 | 0 | 0 | 0 |
|  | NPI-Q | AV45 | 0 | 0 | 0 | 0 | 0 | 0 |
|  |  | PiB | 0 | 0 | 0 | 0 | 0 | 0 |

**Table 9:** Multivariate Linear Regression Analysis-Region of Interest Across Clinical Conditions for Both Threshold Schemes for Eigenvector Centrality

| Clinical Condition |  |  | CN |  | MCI |  | AD |  |
| --- | --- | --- | --- | --- | --- | --- | --- | --- |
| Graph Metric | PT | Tracer | OMST | SPT | OMST | SPT | OMST | SPT |
| Eigenvector Centrality | MMSE | AV45 | 2 | 2 | 11 | 4 | 7 | 10 |
|  |  | PiB | 2 | 2 | 6 | 6 | 8 | 3 |
|  | NPI-Q | AV45 | 1 | 4 | 1 | 1 | 0 | 0 |
|  |  | PiB | 3 | 0 | 3 | 3 | 2 | 9 |

**Table 10:** Multivariate Linear Regression Analysis-Region of Interest Across Clinical Conditions for Both Threshold Schemes for Closeness Centrality

| Clinical Condition |  |  | CN |  | MCI |  | AD |  |
| --- | --- | --- | --- | --- | --- | --- | --- | --- |
| Graph Metric | PT | Tracer | OMST | SPT | OMST | SPT | OMST | SPT |
| Closeness Centrality | MMSE | AV45 | 0 | 5 | 1 | 2 | 2 | 1 |
|  |  | PiB | 2 | 4 | 46 | 0 | 6 | 0 |
|  | NPI-Q | AV45 | 1 | 0 | 0 | 2 | 0 | 0 |
|  |  | PiB | 3 | 30 | 1 | 0 | 0 | 10 |

PT: Psychometric Assessment, OMST: Orthogonal Minimum Spanning Tree, SPT: Shortest Path Thresh- old, MMSE: Mini Mental State Examination, NPIQ: Neuropsychiatric Inventory Questionnaire

### 4.5. Collective Influence Ranking

The collective influence algorithm ranks the ROIs; here, the rank list is generated for the two tracers-AV45 and PiB. When a comparison of the rank is carried out between the clinical groups and tracers in the case of PiB, the ranking increases when moving from CN clinical condition to MCI, and then ranking decreases going from MCI to AD. Overall the ranking increases by 50% from cognitively normal condition to Alzheimer’s disease condition.

### 4.6. Demographics

On the basis of the selection criteria, 531 patients were available for this study. Of this, 48% of the females were of the Cognitively Normal group, 25% with Mild cognitive impairment, and 27% with Alzheimer’s disease.

43% of the patients received more than 12 years of education as opposed to only 16% who received less than 12 years of education, 31% received more than 12 years of education in the MCI group as opposed to 69% with less than 12 years of education.

47% of the Left-handed patients were in the AD clinical group as opposed to 26% in the right-handed patients. One patient in the MCI, two in CN, and four in AD groups were multilingual.

## 5. Discussion

Here, a comparison of five nodal metrics; Betweenness centrality, closeness centrality, current flow betweenness centrality, Eigenvector centrality, and percolation centrality, is carried out to better understand the study. Based on the variance analysis and multivariate regression testing and the percolation centrality graph metric computed using the PET images, it is possible to show Alzheimer’s disease progressing through the beta-amyloid/tau protein networks.

The student t-test provides nodes for each of the five centrality measures across the three clinical conditions, tracer types, and threshold schemes. Here, only the Current Flow Betweenness Centrality Measure fails to provide ROIs across all conditions (see tables 6 through 10), provides only three in cognitively normal condition and two in a mild cognitively impaired condition, both using the OMST scheme for threshold.

It is observed that percolation centrality values of certain areas of the brain, such as inferior and superior parietal lobules, are reliable for the tracer PiB. In contrast, for most other cases, the brain areas differ for each tracer considerably. The variation due to the tracers could be because AV45 and PiB bind to the amyloids differently. It is also observed that the percolation centrality of Broca’s area is a reliable differentiator between C.N. and A.D. clinical conditions, which validates previous findings that cognitive impairment affects speech production[54].

The MLR analysis for each of the centrality measures across the clinical conditions for both the tracer types provides, on average, one to two ROIs across conditions that can be considered as markers for studying Alzheimer’s disease. Expect the current flow betweenness centrality measure, which performed poorly in this study providing only four(3-CN, 1-MCI) ROIs for both the threshold schemes and only one(MCI) ROI for AV45 and PiB tracers, respectively.

Further, when MLR is carried out for the NPI questionnaire, it provides fewer ROIs for each of the centrality measures. When comparing the contribution of ROIs for MMSE-related tasks and NPIQ related tasks. Percolation centrality has the highest percentage of 41% of ROIs. Followed by Closeness centrality with 40.5%, Betweenness centrality with 30%.

The results from the Scheffe test provide a means to validate and increase the confidence in the results—the leave one out cross-validation (LOOCV) strategy to test the robustness and reliability of our regression. Here cross-validation strategy is implemented because it is unbiased and better suited to our smaller sample size. Using the regularization (L1 - Lasso or L2 - Ridge) to control for overfitting, it is observed that increasing regularization on validation RMSE.

The ROIs obtained from the pairwise t-test for between the clinical conditions show that the OMST scheme provides a higher number of valid ROIs across the five centrality measures. In comparison between the two tracer types and threshold schemes, AV45 provides 92 ROIs, whereas PiB gives 88 ROIs. Of this, 33.7% and 39.8% of the ROIs are based on the Shortest Path threshold scheme.

Previous studies show that the seeding of amyloid-beta occurs in neocortical and subcortical regions[55]; from this study, it is observed that for PiB, the following ROI - W.M. Superior occipito-frontal fascicle R is part of both the neocortical and subcortical regions of the brain. Apart from this, AV45 tracer has G.M. Medial geniculate body L ROI in the subcortical region and the following in the neocortical region -G.M. Superior parietal lobule 7P L, G.M. Anterior intra-parietal sulcus hIP3 R, G.M. Superior parietal lobule 7A L, and G.M. Superior parietal lobule 5L L, these are picked up with OMST scheme across all conditions when compared to SPT.

Prior research shows that damage to the parietal lobe is common in A.D., which can lead to apraxia[54,56], which is attested by these results. A.D. is associated with atrophy of the cornu ammonis, the subfield of the hippocampus, and deficits in episodic memory and spatial orientation[57–59]

And the following in the subcortical region - GM Amygdala-laterobasal group L, GM Amygdala-laterobasal group R and GM Hippocampus hippocampal-amygdaloid transition area R. Age factor not so important but the presence of beta amyloid deposits is[60], Since these ROIs stand out irrespective of the clinical condition or demographic backgrounds, the percolation centrality has a potential to be a reliable value for AD diagnosis, these ROIs are picked up by both tracers and threshold schemes.

Recent methods include genetic and protein markers to improve predicting the course of the disease[61], Genetic testing[62]for markers of A.D., the apolipoprotein-e4 (APOE-e4)[63], or the use of blood testing or brain imaging to rule out dementia due to other factors. These methods rely on many data points and equally reliable computing hardware; this is currently a challenge.

Given that A.D. diagnosis is a global challenge, a method that works well in a spectrum of nations, from developed countries such as the United States to rural hospitals of southeast Asia or Africa[2] is necessary. Methods such as principal component analysis have a few drawbacks; for instance, choosing the number of principal components and data standardization for multiple PET scans of patients with different tracers leads to controlling multiple variables. Or using regression analysis which is based on the assumption that there are cause and effect in place. Furthermore, a relationship that is present within a limited data set might get overturned with a detailed data set.

### 5.0.1. Limitations

This study does not give any evidence regarding the disease progression in terms of the ROIs or patient clinical group. However, this can be addressed by increasing the number of observations within each patient clinical group.

The PET tracers used for acquiring the images, Pittsburgh Compound B (PiB) and Florbetapir (AV45), are compared to check for which among the two tracers provide a more consistent or reliable PCv. Here, the AV45 tracer binds with a high affinity to the beta-amyloid plaque, whereas PiB binds to oligomers or protofibrils. A possible explanation for the difference in PCv generated using these tracers would be their binding targets. The use of second-generation tracers can help improve the accuracy and test the applicability of percolation centrality on other neurodegenerative diseases and the possibility of using it in metastatic cancer scenarios.

Expanding the dataset to include more patients and comprehensive data that factors in healthy aging shrinkage of the brain, which results in a decrease of the distances of the brain networks, can help improve the reliability of the percolation centrality value. This can then provide a setting for testing out other psychological assessments that can be used as early indicators for dementia due to Alzheimer’s disease, thereby tailoring it to specific demographics or population subsets.

The current pipeline is built for tracers such as AV45 and PiB, which indicate beta-amyloid plaque concentrations directly and as a post-hoc implementation. However, the pipeline can work with second-generation tracers and tracers like FDG with some appropriate modifications, namely: taking the multiplicative inverse of the percolation states of each of the ROIs to reflect the behavior of the FDG tracer.

A comparison of the ROIs across clinical conditions and tracers does not provide any new information at this stage regarding a common or group of common ROIs across the data(see Table 6. and 7.).

## 6. Conclusions

This study shows that percolation centrality is a reliable predictor and identifies the nodes that regulate the movement of beta-amyloid plaque and use them to track the disease.

This work demonstrates that using the existing neuroimaging method, PET-CT, can add value with relatively short computation time provided sufficient hardware capability is present. The ability to provide a metric to the extent of the disease state is advantageous to the current world of Alzheimer’s. Prolonging life with modern-day medicine pushes patients to a world of medical experiences that deviate from the normal. Being able to show the deviation with a value such as percolation centrality has potential applications.

The reliability of percolation centrality can be improved by addressing the concerns that arise by the factors such as the number of patients and the number of patients within each clinical group, time points of data collection, demographics, and the PET tracers used were the limiting factors. Thus, this study provides the usability of percolation centrality value to determine the patient’s state and sets the stage for studying other neurodegenerative diseases.

Unlike measures such as hub centrality or betweenness centrality, which provide information regarding a vital vertex/node within a network, the collective influence algorithm provides a minimum set of nodes of the network that are key to the beta-amyloid plaque movement, which can provide information regarding a particular pathway that is susceptible to the neuropathology.

The threshold schemes implemented in this study indicate that a data-driven approach such as the Orthogonal Minimum Spanning Tree provides better results compared to the Shortest Path approach. Finally, we rank the ROIs based on influence in the network using the CI algorithm. We compare the results on two thresholding approaches, SPT and OMST, along with finding the influential nodes; this will help us gauge the reliability across different threshold schemes. CI algorithm gives a ranked list of influential nodes for each network (or each scan).

The rank of a node is then further calculated for each category as the sum of individuals ranks of that node for every scan-wise list, divided by the number of scans it occurs in. Nodes are then ranked accordingly in a given category. This provides a general ranking of nodes in a category (AD/MCI/CN) instead of looking at influential nodes in each scan separately. The results differ slightly based on the thresholding scheme adopted but broadly align with MLR results discussed earlier. Since this is an exploratory study, improving robustness across different thresholding schemes can be a possible future work.

## Data Availability

PET image data is obtained from the Alzheimer's Disease Neuroimaging Initiative. All processed data will be provided on request.

http://adni.loni.usc.edu

## Citation

Kumar B., Gautam.; Prasad, Raghav.; Mahajan, Pranav.; Baths, Veeky. Percolation Centrality and Collective Influence on Beta-amyloid plaques network., *Journal Not Specified* **2021**, 1, 0. https://doi.org/

## Supplementary Materials

Please follow the link https://www.mdpi.com//1/1/0/https://github.com/raghavprasad13/ADNI-Project for the analysis pipeline code, and follow this link https://www.mdpi.com//1/1/0/https://drive.google.com/file/d/1ZIVb6TFyJt68wb_N8mWgBr3xuhOcdZem/view?usp=sharing for Tables 11 through 18.

## Author Contributions

Conceptualization, Gautam and Veeky; methodology, Raghav and Gautam; validation, Gautam, Raghav and Pranav; formal analysis, Gautam, Raghav and Pranav; resources, Veeky Baths; data curation, Gautam and Raghav; writing—original draft preparation, Gautam; writing—review and editing, Gautam, Raghav, Pranav and Veeky; visualization, Raghav and Gautam; supervision, Veeky Baths; All authors have read and agreed to the published version of the manuscript.

## Institutional Review Board Statement

Institutional Review Board Waiver Statement-Informed consent from the patients is obtained before the assessment carried out by the ADNI study team (See ADNI website for details), and this study is a secondary data analysis of the ADNI data collection, which aims at providing a simplified metric to an already diagnosed patient. The data access and usage is within the ADNI data use agreements.

## Informed Consent Statement

Informed consent was obtained from all patients involved in the study by ADNI study Team(s).

## Data Availability Statement

PET images from ADNI database are used in this study.

## Abbreviations

The following abbreviations are used in this manuscript:

AD: Alzheimer Disease
ANOVA: Analysis of Variance
AV45: Florbetapir (18F-AV-45)
CI: Collective Influence
CN: Cognitively Normal
CSF: Cerebrospinal Fluid
DOF: Degrees of Freedom
EEG: Electroencephalography
FAB: Frontal Assessment Battery
fMRI: functional Magnetic Resonance Imaging
FSL: FMRIB Software Library
GNU: GNU’s Not Unix
PCv: Percolation Centrality Value
PET: Positron Emission tomography
PiB: Pittsburgh compound B (11C-PIB)
RMSE: Root Mean Square Error
MCI: Mild Cognitive Impairment
MEG: Magnetoencephalography
MLR: Multivariate Linear Regression
MMSE: Mini-Mental State Examination
MST: Minimum Spanning Tree
NPIQ: Neuropsychiatric Inventory Questionnaire
OMST: Orthogonal Minimum Spanning Tree

## Appendix A.

### Appendix A.1. Percolation Centrality Computation

The percolation centrality value is calculated for each network using the inbuilt function of Networkx. This has a worst-case time complexity of O(n^3^), where n is the number of nodes in the network. Using a modified form of Brandes’ fast algorithm for betweenness centrality, the complexity can be reduced to O(nm), where m is the number of edges. However, percolation centrality calculation with target nodes cannot take advantage of this optimization and has a worst-case time complexity of O(n^3^)

### Appendix A.2. Other Graph Metrics-Tables

